# A report-supervised EEG-language foundation model enables broad, interactive clinical interpretation

**DOI:** 10.64898/2026.11.18.26368408

**Authors:** Dominik D. Kranz, Robert Terziev, Michał Łukomski, Laura Krumm, Christian Meisel

**Affiliations:** Section on Computational Neurology, Berlin Institute of Health and Department of Neurology, Charité – Universitätsmedizin Berlin

**Author notes:** Shared first authorship (Dominik D. Kranz, Robert Terziev).

## Abstract

Medical AI for complex signals such as electroencephalography (EEG) relies on costly expert annotations and predefined diagnostic labels that fail to capture the richness and nuance of clinical interpretation. Whether models trained on routine clinical reports can overcome these limitations while matching expert-supervised performance remains unclear. We introduce EEG Copilot, a biosignal–language foundation model trained on routine EEG–report pairs without dedicated task-specific expert annotations. Across two external datasets (6,078 EEGs), EEG Copilot performed on par with a state-of-the-art expert-supervised model across 11 shared findings (mean AUROC 0.80–0.81), corroborated by direct neurologist review of 200 EEGs. Beyond predefined diagnostic categories in conventional EEG AI, EEG Copilot assessed 23 clinical characteristics through natural-language questions in English and German across 9,689 EEGs from four hospitals (mean AUROC 0.80–0.82), demonstrating that automated EEG assessment need no longer be confined to fixed diagnostic outputs. Crucially, EEG Copilot generated integrated clinical impressions directly from EEG rather than isolated predictions with shuffled-report controls supporting recording-specific content and neurologist review of 800 impressions showing that most were accurate or clinically usable with minor errors. These findings chart a path beyond label-bound medical AI, transforming routine clinical reports into scalable supervision for flexible, language-driven assessment and integrated interpretation of complex physiological signals.

## Introduction

Much of medical artificial intelligence (AI) follows a common paradigm: medical experts annotate datasets for specific diagnostic tasks, and models are trained to predict the resulting labels. Creating these dedicated annotations is labor-intensive, expensive and difficult to scale. More fundamentally, in many clinical settings, predefined labels capture only part of clinical interpretation, whereas physicians must integrate multiple findings, assess their relationships and clinical significance, and communicate nuanced conclusions in medical language. This paradigm thus constrains not only how medical AI is trained, but also the complexity and flexibility of the interpretations it can provide.

Electroencephalography (EEG) exemplifies these challenges. A cornerstone of epilepsy diagnosis, the assessment of cerebral dysfunction and neurological monitoring in critically ill patients, EEG interpretation is among the most demanding tasks in routine clinical diagnostics. Recordings comprise high-dimensional, temporally dynamic and artifact-prone signals, in which clinically decisive information may be subtle, transient and distributed across multiple channels and timescales. Accurate interpretation requires integrating waveform morphology, frequency, spatial distribution, temporal evolution and physiological context into a clinical clinical narrative. Yet this complex and labor-intensive process depends on specialized expertise that is unevenly available, with potentially serious consequences of misinterpretation for diagnosis and treatment. AI thus holds considerable promise for making specialized EEG expertise more widely accessible while improving the efficiency and consistency of clinical interpretation.

Automated EEG analysis has made substantial progress, with supervised learning approaches achieving high diagnostic performance across a range of predefined EEG findings^1,2^. However, these advances have depended on extensive task-specific annotations by EEG experts, with each additional finding or finer diagnostic distinction potentially requiring further costly labeling efforts. More fundamentally, predefined output categories restrict what these systems can explicitly communicate, whereas clinical EEG reports capture gradations, relationships, uncertainty and context-dependent interpretations that extend beyond fixed diagnostic labels. Such rich narrative reports are generated routinely during clinical care yet remain largely untapped as a source of supervision for developing more scalable and flexible EEG AI.

Recent advances in self-supervised and multimodal foundation models suggest a fundamentally different approach to clinical EEG interpretation. Rather than training separate classifiers on manually curated labels, models could learn directly from the vast collections of EEG recordings and corresponding clinical reports generated during routine care. In this paradigm, medical language would serve not merely as an output format, but as a source of supervision through which models learn to represent complex physiological information, its relationships and clinical meaning. Such models could support a broad and extensible repertoire of clinical findings, enabling natural-language questions, contextual interpretation and narrative report generation without requiring dedicated annotations or classifiers for each task. Related approaches have been explored in computational pathology and EEG–language^3,4^ modeling. This raises a fundamental question: can heterogeneous and imperfect clinical reports provide sufficiently rich supervision for a new generation of medical AI—one that learns from routine care and interprets complex physiological signals beyond predefined diagnostic categories?

Here we introduce EEG Copilot, a biosignal–language foundation model that learns from routinely acquired EEG recordings and clinical reports without dedicated task-specific expert annotations. We demonstrate that EEG Copilot achieves diagnostic performance on par with a state-of-the-art expert-supervised EEG foundation model across shared clinical findings, while supporting a substantially broader range of EEG characteristics. Beyond predefined classifications, the model enables natural-language questioning and generates integrated clinical impressions that describe and contextualize EEG findings. Together, these findings demonstrate a fundamental shift in how medical AI can be developed and applied - combining scalable learning from routine clinical data with state-of-the-art diagnostic performance and flexible, language-based interpretation of complex physiological signals.

## Results

### Learning clinical EEG interpretation from routine recordings and reports

We developed EEG Copilot to establish whether learning directly from routine EEG recordings and clinical reports can achieve diagnostic performance comparable to expert-supervised AI, while enabling a broader repertoire of clinical findings and nuanced interpretation through natural language (Fig. 1). EEG Copilot processes multichannel EEG recordings of up to 2 h and supports natural-language questioning, follow-up dialogue and generation of clinical impressions in English and German (Fig. 1a). The architecture combines a self-supervised EEG encoder with a hierarchical EEG– language bridge and a large language model, linking physiological signals to clinical language within a unified framework (Fig. 1b). Model development used routinely acquired EEG recordings and accompanying clinical reports from Massachusetts General Hospital (MGH) and Brigham and Women’s Hospital (BWH)^5^.

**Figure 1.**
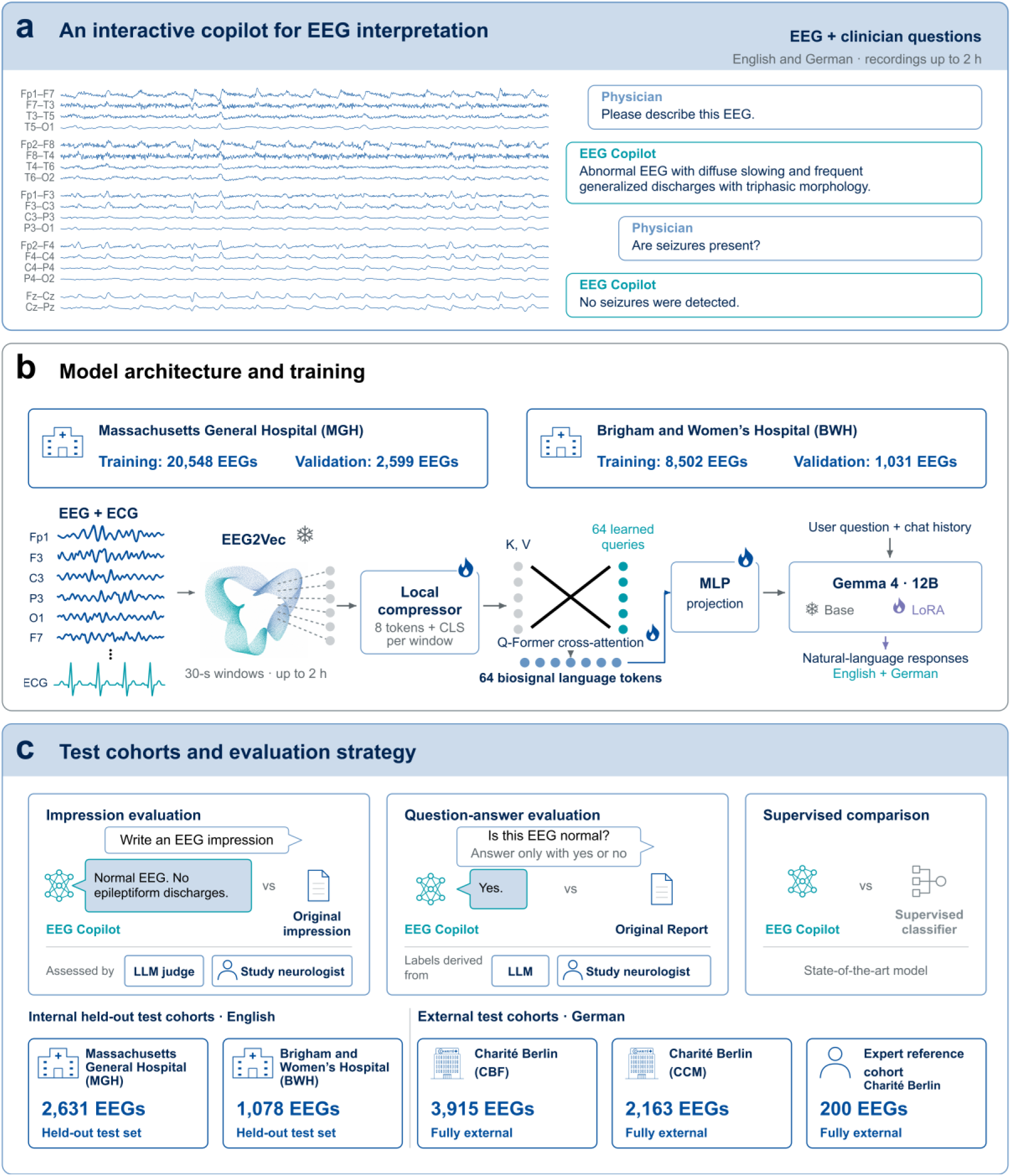
EEG Copilot architecture and evaluation framework. **a,** Illustrative interaction with EEG Copilot, enabling natural-language questioning, follow-up dialogue and generation of clinical impressions from multichannel EEG recordings of up to 2 h, in English and German. **b,** Model architecture and development cohorts. A self-supervised EEG encoder (EEG2Vec) processes 30-s EEG and electrocardiography (ECG) windows. A hierarchical EEG–language bridge aggregates recording-level information into c4 tokens, which are projected into a large language model (Gemma 4 12B) to generate responses conditioned on EEG signals, clinician questions and conversation history. Training used routinely acquired EEG recordings and clinical reports from Massachusetts General Hospital (MGH; 20,548 training, 2,5SS validation) and Brigham and Women’s Hospital (BWH; 8,502 training, 1,031 validation). Snowflakes and flames denote frozen and trainable components. **c,** Evaluation framework spanning diagnostic question answering, clinical impression generation and comparison with an expert-supervised EEG model. Evaluation included held-out MGH (n = 2,c31) and BWH (n = 1,078) cohorts, external Charité cohorts (CBF, n = 3,S15; CCM, n = 2,1c3) and direct neurologist review of 200 EEG recordings. Diagnostic findings were evaluated against report-derived or expert EEG-based references; generated impressions were assessed by an LLM judge and neurologist review. Cohort sizes are shown; analysis-specific denominators vary. MLP, multilayer perceptron; LoRA, low-rank adaptation^c^ .

We evaluated EEG Copilot in held-out cohorts from both development hospitals and two external Charité campuses in Berlin (Fig. 1c). The evaluation examined diagnostic discrimination across diverse clinical EEG findings, performance against a state-of-the-art expert-supervised EEG foundation model, and the quality of generated clinical impressions. In addition to large-scale evaluations against report-derived references, direct neurologist review of 200 EEG recordings provided an independent reference based on the underlying signals.

To learn clinical EEG interpretation without dedicated task-specific annotations, we developed a three-stage training strategy (Fig. 2). First, we developed EEG2Vec, a self-supervised encoder pretrained on 2.28 million 30-s EEG windows without clinical labels (Fig. 2a–c). Second, a hierarchical EEG–language bridge compressed representations across entire recordings into 64 tokens and learned their alignment with corresponding clinical reports (Fig. 2d). Finally, we adapted a large language model using report-derived impressions, clinical questions and multi-turn dialogues, enabling flexible EEG-conditioned interpretation without dedicated task-specific expert annotations.

**Figure 2.**
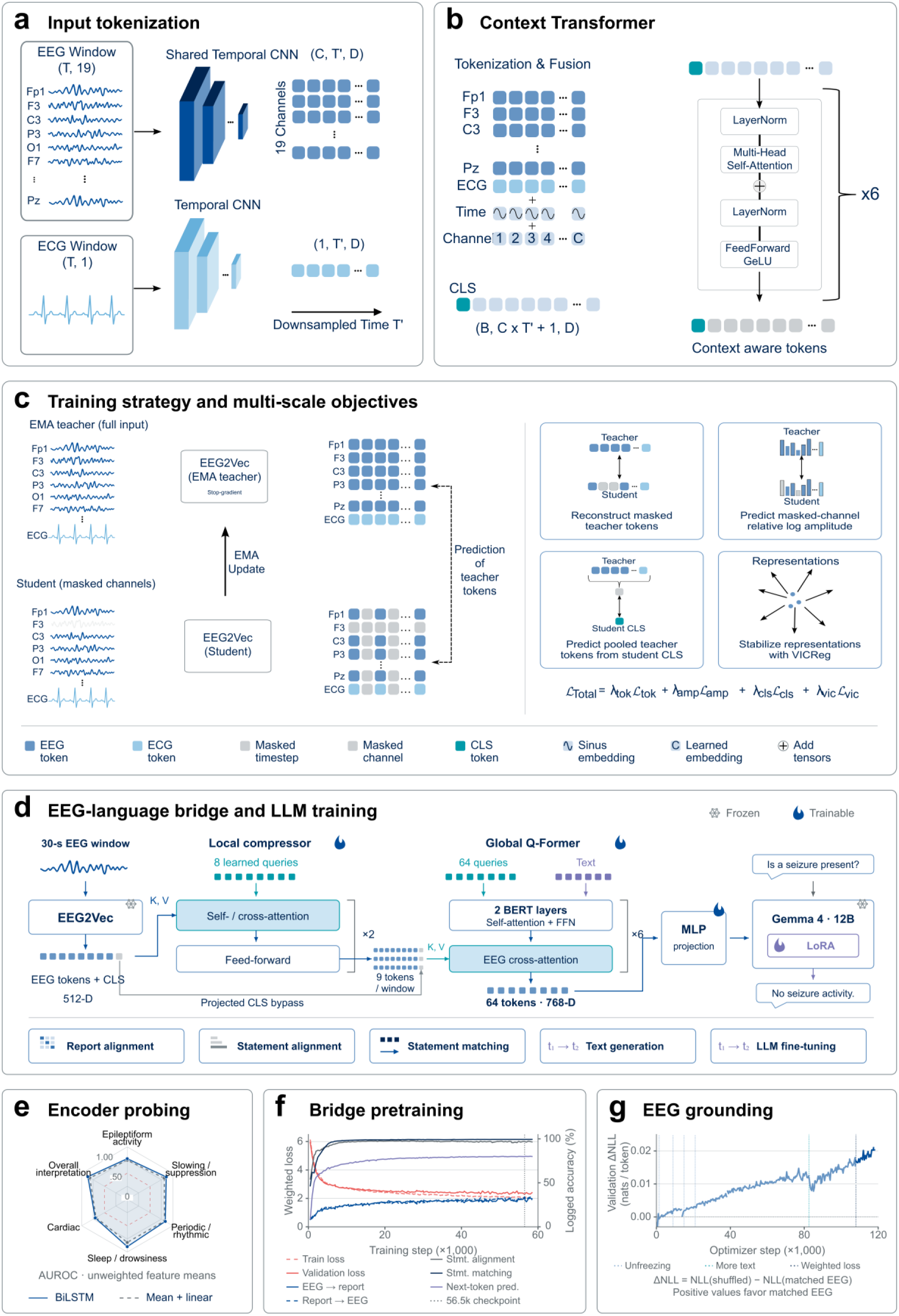
Self-supervised EEG representation learning and clinical language alignment. **a,** Temporal convolutional networks tokenize 1S EEG channels and one electrocardiography (ECG) channel into 30-s window representations. **b,** A six-layer context transformer^7^ integrates EEG and ECG tokens with temporal and channel embeddings and a summary (CLS) token. **c,** Self-supervised EEG2Vec pretraining. A student network learns from channel-masked inputs by predicting representations generated from complete recordings by an exponential-moving-average (EMA) teacher. Training combines masked-token prediction, channel-amplitude prediction, pooled-representation prediction and variance–invariance–covariance regularization (VICReg^8^). **d,** Hierarchical EEG– language alignment. A local compressor produces nine tokens per window, which a recording-level query transformer aggregates into c4 tokens. Training aligns EEG representations with clinical reports and individual statements, followed by text generation and language-model adaptation using low-rank adaptation (LoRA). **e,** Clinical information captured by frozen EEG2Vec representations, assessed using bidirectional long short-term memory (BiLSTM^S^) and linear probes on 3,380 held-out recordings. Radar plots show mean area under the receiver operating characteristic curve (AUROC) across clinical categories, averaged over three training seeds. **f,** EEG–language bridge pretraining, showing training and validation losses and task-specific validation accuracies. The vertical line marks the selected checkpoint. **g,** EEG grounding assessed by the difference in reference-response negative log-likelihood between shuffled and matched EEG inputs (ΔNLL). Positive values indicate greater response likelihood with the corresponding EEG recording. Dotted lines indicate training-stage changes; the trajectory is descriptive because evaluation settings varied across restarts.

Self-supervised pretraining yielded representations containing substantial clinically relevant information. Across 3,380 held-out recordings, supervised probes trained on report-derived labels discriminated 24 EEG and ECG findings, achieving mean areas under the receiver operating characteristic curve (AUROCs) of 0.841 with recurrent readouts and 0.804 with linear readouts (Fig. 2e; Supplementary Fig. 1). Subsequent EEG–language training established cross-modal alignment (Fig. 2f), with matched EEG inputs yielding higher reference-response likelihoods than shuffled recordings (Fig. 2g), indicating that language predictions incorporated recording-specific information.

### EEG Copilot achieves diagnostic performance on par with expert-supervised EEG AI

To determine whether learning from routine clinical reports can achieve diagnostic performance comparable to conventional expert-supervised approaches, we directly benchmarked EEG Copilot against MORGOTH, a state-of-the-art EEG foundation model developed using extensive expert-annotated datasets^2^. We evaluated both models on 11 shared clinical findings across 6,078 EEG recordings from two external Charité cohorts (CCM, n = 2,163; CBF, n = 3,915), using the same recordings and report-derived reference labels for each finding (Fig. 3a). Unlike MORGOTH, EEG Copilot had not received dedicated expert annotations for any of these diagnostic tasks.

**Figure 3.**
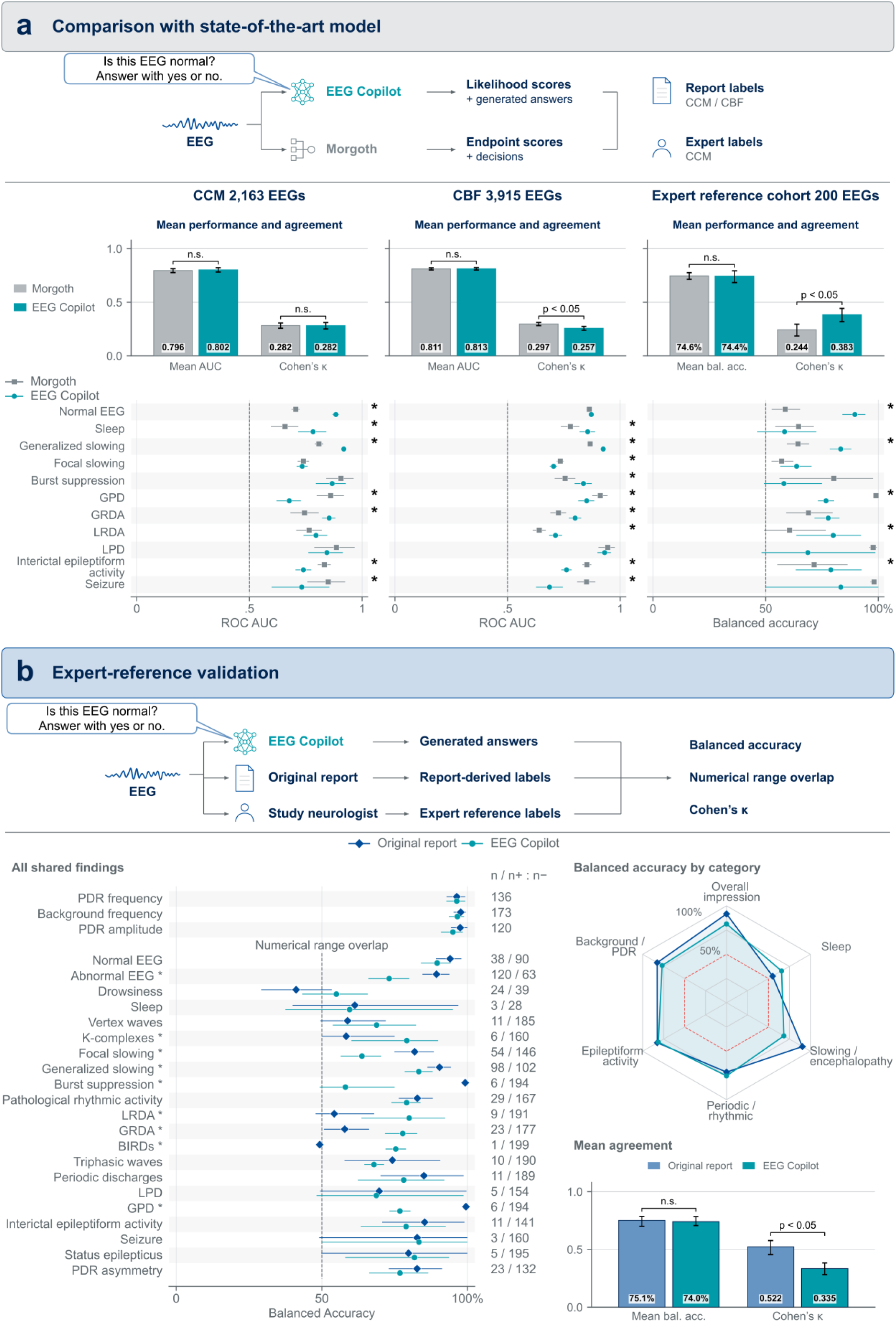
Comparison with expert-supervised EEG AI and validation against direct neurologist review. **a**, Comparison of EEG Copilot and MORGOTH across 11 shared clinical findings. Both models were evaluated on the same assessable recordings and reference labels for each finding. In the external CCM (n = 2,1c3) and CBF (n = 3,S15) cohorts, discrimination was assessed using continuous-score AUROC against report-derived labels; in the direct expert-reference cohort (n = 200), binary decisions were assessed using balanced accuracy against neurologist labels. EEG Copilot provided answer likelihood scores and generated responses, whereas MORGOTH provided finding-specific scores and decisions. Upper panels show unweighted mean AUROC or balanced accuracy across the 11 findings and mean Cohen’s κ across findings with at least ten positive and ten negative references; lower panels show finding-level AUROC or balanced accuracy with S5% confidence intervals. Unmentioned non-sleep findings in clinical reports were treated as negative, whereas unmentioned sleep findings were excluded as unknown. MORGOTH sleep scores were derived from the maximum of its N1 and N2 recording-level scores, using a threshold of 0.5 for binary decisions. **b**, Expert-reference validation of EEG Copilot and the original clinical reports against direct neurologist review of 200 EEG recordings. The three numerical findings are assessed by overlap between predicted and expert-reference ranges; 21 evaluable binary findings are assessed using balanced accuracy, defined as the mean of sensitivity and specificity. Counts indicate assessable recordings for numerical findings and reference-positive/reference-negative counts for binary findings. The radar plot summarizes balanced accuracy by clinical category; the overall mean includes 21 binary findings. Mean Cohen’s κ summarizes the 12 findings with at least ten positive and ten negative expert-reference cases. Error bars indicate percentile S5% confidence intervals from 5,000 paired recording-level bootstrap resamples. Brackets indicate paired comparisons (n.s., not significant; p < 0.05 where the S5% interval for the paired difference excludes zero); asterisks beside the finding-level comparisons in a indicate paired model-difference intervals excluding zero. Comparisons are exploratory and unadjusted for multiplicity. PDR, posterior dominant rhythm; GPD/LPD, generalized/lateralized periodic discharges; GRDA/LRDA, generalized/lateralized rhythmic delta activity; BIRDs, brief potentially ictal rhythmic discharges.

Despite these fundamentally different supervision strategies, EEG Copilot achieved virtually identical aggregate diagnostic discrimination to MORGOTH, with mean AUROCs of 0.802 versus 0.796 at CCM (p = 0.543) and 0.813 versus 0.811 (p = 0.882) at CBF, respectively (two-sided paired recording-level bootstrap tests; Fig. 3a). Performance varied across individual findings, with neither model consistently outperforming the other. Thus, learning directly from routine EEG–report pairs yielded diagnostic performance on par with an extensively expert-supervised model across the evaluated shared tasks.

Because routine clinical reports are themselves imperfect references, we additionally evaluated both models against an independent assessment based on direct neurologist review of 200 EEG recordings. Against this EEG-based expert reference, mean balanced accuracy across the 11 shared findings was 0.744 for EEG Copilot and 0.746 for MORGOTH (p = 0.940; Fig. 3a). Cohen’s κ provided a complementary perspective on agreement: mean κ was identical for both models at CCM (0.282; p = 0.979), lower for EEG Copilot at CBF (0.257 versus 0.297 for MORGOTH; p = 0.0002), but higher for EEG Copilot against the direct expert reference (0.383 versus 0.244; p = 0.0004). All p values were calculated using two-sided, null-centered paired whole recording bootstrap tests. These results independently corroborate the comparable diagnostic performance observed across the larger external cohorts.

Direct neurologist review also enabled comparison of EEG Copilot with the original clinical reports (Fig. 3b). Across 21 clinical findings, mean balanced accuracy was similar for EEG Copilot and the original reports (0.740 versus 0.751; P = 0.667), although the original reports showed higher agreement with the expert reference as measured by Cohen’s κ across 12 eligible findings (0.522 versus 0.335; P = 0.0002). Both comparisons used two-sided, null-centered paired whole-recording bootstrap tests. Together, these findings demonstrate that competitive diagnostic performance can be achieved without extensive task-specific expert annotation, establishing routine clinical documentation as a scalable source of supervision for medical AI.

### Natural-language questioning extends diagnostic assessment across diverse EEG findings

Beyond achieving diagnostic performance on par with expert-supervised EEG AI on shared tasks, a central advantage of EEG Copilot is its ability to assess a substantially broader repertoire of clinical findings through natural-language questions in both English and German. Unlike conventional classification-based approaches, EEG Copilot does not require a predefined output head for each clinical finding, allowing diverse EEG characteristics to be assessed through a single language-conditioned model without additional task-specific training. We evaluated 23 EEG characteristics spanning overall interpretation, sleep, slowing and encephalopathy, periodic and rhythmic patterns, and epileptiform activity (Fig. 4), extending its evaluated diagnostic repertoire well beyond the 11 findings available for direct comparison with MORGOTH^2^.

**Figure 4.**
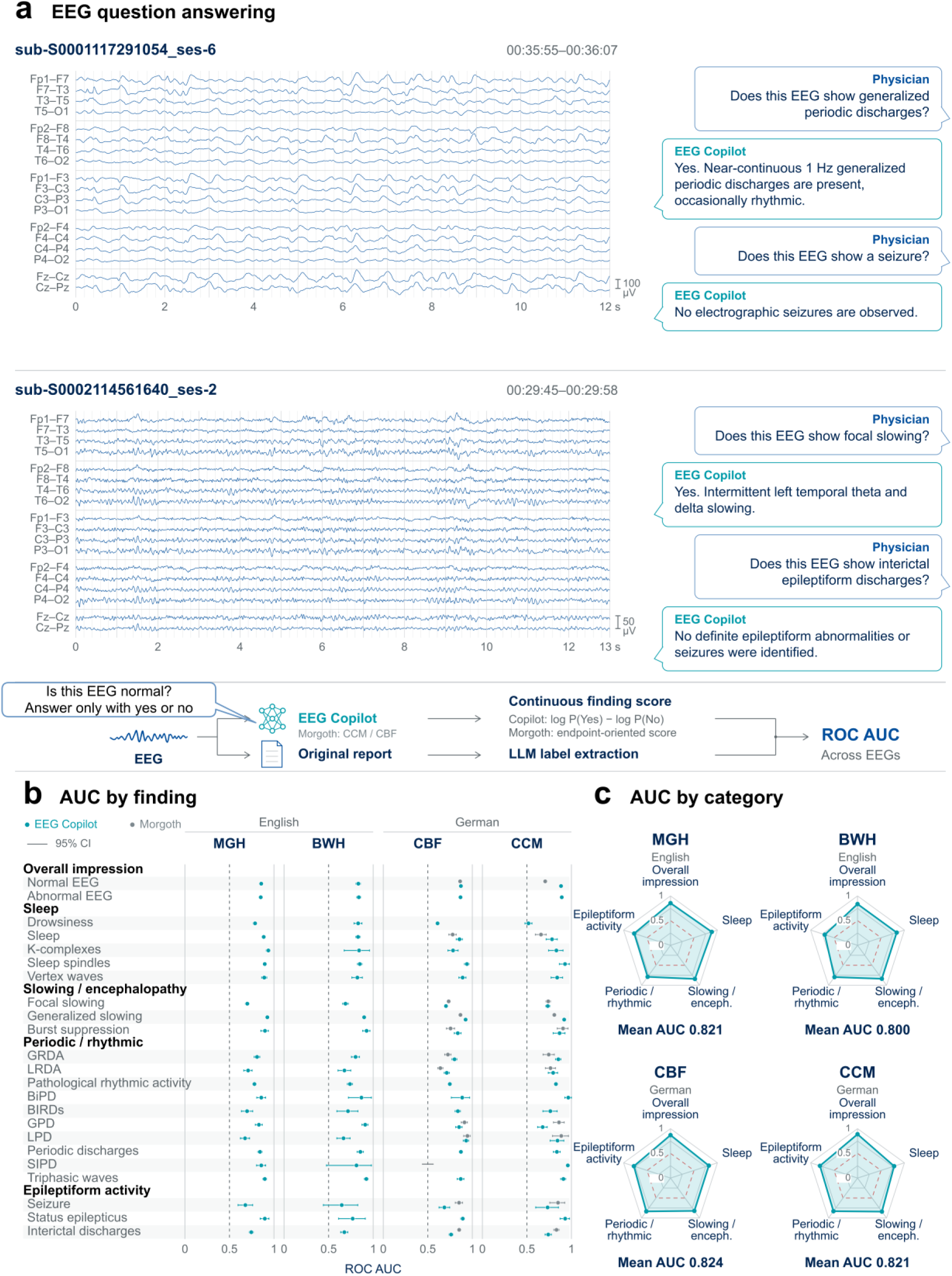
Multilingual assessment of diverse clinical EEG findings through natural-language questioning. **a,** Example EEG recordings with targeted clinical questions and EEG Copilot responses, followed by the evaluation workflow. Continuous finding scores are calculated from the difference in affirmative and negative answer log-likelihoods (log P(Yes) − log P(No), or log P(Ja) − log P(Nein) in German) and evaluated against report-derived reference labels. **b,** Finding-level areas under the receiver operating characteristic curve (AUROCs) across four cohorts: MGH (n = 2,557), BWH (n = 1,054), CBF (n = 3,S15) and CCM (n = 2,1c3). Ǫuestions were posed in English for MGH/BWH and German for CBF/CCM. Teal points represent EEG Copilot; grey points represent MORGOTH on shared findings in the external cohorts. Horizontal lines indicate S5% confidence intervals from 5,000 recording-level bootstrap resamples. Unmentioned non-sleep findings were treated as negative; unmentioned sleep findings and other unknown labels were excluded. **c,** Mean AUROCs across five clinical categories, demonstrating diagnostic discrimination across cohorts and languages. Overall means include the same 22 evaluable findings at all sites; sharp-and-slow-wave periodic discharges (SIPD), lacking positive references at CBF, were excluded from category and overall means. Dashed lines indicate chance performance (AUROC = 0.5). BiPD, bilateral independent periodic discharges; BIRDs, brief potentially ictal rhythmic discharges; GPD/LPD, generalized/lateralized periodic discharges; GRDA/LRDA, generalized/lateralized rhythmic delta activity.

We evaluated this capability across 9,689 EEG recordings from four hospital cohorts in the United States and Germany, using English questions for the Boston cohorts and German questions for the Charité cohorts. Each clinical finding was assessed using the difference in likelihood between affirmative and negative answers to a corresponding natural-language question, with discrimination evaluated against findings extracted from routine clinical reports (Fig. 4a). Across the 22 findings evaluable at all four sites, mean AUROC was remarkably consistent: 0.821 at MGH (95% CI, 0.810–0.831), 0.800 at BWH (0.778–0.822), 0.824 at CBF (0.814–0.834) and 0.821 at CCM (0.805–0.837; Fig. 4b,c). Thus, EEG Copilot maintained comparable aggregate diagnostic discrimination across English-and German-language clinical settings, despite differences in patient populations and reporting practices, without site-specific retraining.

Diagnostic performance was particularly strong for generalized slowing (AUROC 0.898– 0.926), triphasic waves (0.870–0.923) and sleep spindles (0.849–0.940), while discrimination was more modest for transient abnormalities such as interictal epileptiform discharges (0.676–0.762) and seizures (0.647–0.732; Fig. 4b). The examples in Fig. 4a further illustrate how targeted questions can elicit descriptions of EEG abnormalities, including their morphology and spatial distribution. Together, these findings establish a central advantage of EEG–language modeling: a broad and extensible repertoire of clinical EEG characteristics accessible through a single multilingual, natural-language interface, without dedicated expert annotations or separate classifiers for each finding.

### EEG Copilot generates integrated clinical interpretations in natural language

Beyond identifying individual EEG findings, EEG Copilot can synthesize multiple observations into comprehensive clinical impressions expressed in natural language. We evaluated this capability by assessing its ability to generate EEG reports describing background activity, sleep, pathological abnormalities and their clinical significance directly from the recordings (Fig. 5a). We assessed these generated impressions across four hospital cohorts using automated evaluations at scale and independent neurologist assessments of their clinical content and usability.

**Figure 5.**
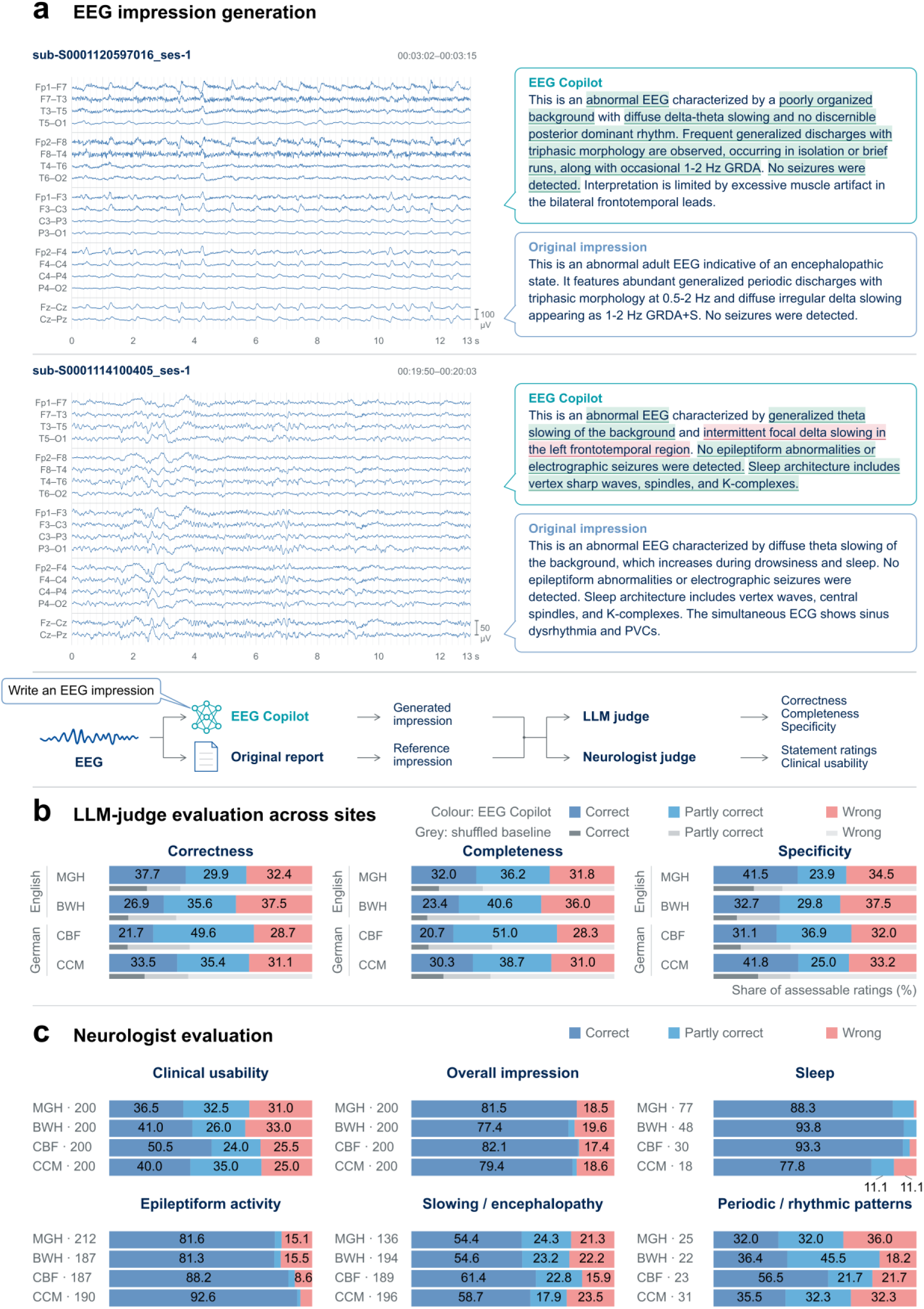
Generation and clinical evaluation of integrated EEG impressions. **a,** Example EEG recordings with corresponding EEG Copilot-generated and original clinical impressions. Highlighted passages indicate neurologist annotations of individual statements. The evaluation combines automated assessment of correctness, completeness and specificity with neurologist assessment of statement accuracy and clinical usability. **b,** Large language model (LLM)-judge evaluation of S,332 generated impressions across four cohorts: MGH (n = 2,378), BWH (n = S73), CBF (n = 3,830) and CCM (n = 2,151). Impressions were evaluated in English (MGH/BWH) or German (CBF/CCM) against their corresponding clinical reports using three-point rating scales. Colored bars show matched evaluations; grey bars show ratings after randomly pairing generated impressions with unrelated reference impressions within each cohort (mean of five permutations). **c,** Neurologist evaluation of 800 generated impressions (200 per cohort). Overall clinical usability was categorized as correct, usable with minor errors or clinically unusable. Individual statements were independently rated as correct, partly correct or wrong across five clinical categories. Neurologist assessments were based on impression texts rather than direct review of the EEG signals. Row labels indicate impression or statement counts; statement-level denominators exclude omitted findings. GRDA, generalized rhythmic delta activity; ECG, electrocardiogram; PVCs, premature ventricular contractions.

Across 9,332 EEG recordings from the United States and Germany, a language-model judge evaluated generated impressions against the corresponding clinical reports for correctness, completeness and specificity, following a recently proposed strategy for automatic report judging^10^ (Fig. 5b). Overall, impressions received full or partial credit for correctness in 67.6% of cases at MGH (95% CI, 65.7–69.5%), 62.5% at BWH (59.4–65.5%), 71.3% at CBF (69.9–72.7%) and 68.9% at CCM (67.0–70.8%). To determine whether this agreement reflected recording-specific information rather than generic clinical descriptions, we repeated the evaluation with randomly shuffled pairings of generated and reference impressions. Correctness ratings declined substantially to 33.0–39.7%, with similar differences in completeness and specificity across all four cohorts. These results demonstrate that EEG Copilot generates clinically differentiated impressions that show substantially greater agreement with their corresponding reports than with unrelated clinical documentation.

An EEG-specialized neurologist additionally evaluated 800 generated impressions, comprising 200 from each hospital cohort (Fig. 5c). Impressions were judged clinically correct or usable with minor errors in 69.0% of cases at MGH (95% CI, 62.5–75.5%), 67.0% at BWH (60.5–73.5%), 74.5% at CBF (68.0–80.5%) and 75.0% at CCM (69.0–81.0%). Statement-level assessment showed high correctness for overall interpretations (range across sites, 77.4–82.1%), sleep-related findings (77.8–93.8%) and epileptiform abnormalities (81.3–92.6%), whereas performance was lower for slowing and periodic or rhythmic patterns. These findings demonstrate the potential of EEG Copilot to generate clinically meaningful narrative interpretations across diverse clinical settings, while identifying areas for further improvement in reporting accuracy.

Together, these results extend the demonstrated capabilities of EEG Copilot from competitive diagnostic classification to the generation of integrated clinical interpretations, showing how a single model trained from routine clinical documentation can support a richer, language-based approach to complex physiological data.

## Discussion

In this study, we demonstrate that a single EEG–language foundation model can achieve diagnostic performance on par with state-of-the-art expert-supervised EEG AI while fundamentally expanding how complex physiological signals can be assessed and interpreted. Trained on routinely acquired EEG recordings and clinical reports without dedicated task-specific expert annotations, EEG Copilot enables assessment of a substantially broader repertoire of clinical EEG characteristics through natural-language questions in English and German, without requiring separate classifiers for each finding. Beyond predefined diagnostic categories, the model generates integrated clinical impressions, bringing automated EEG interpretation closer to the way physicians describe, contextualize and communicate findings in clinical practice. Together, these advances establish a new paradigm for medical AI in which clinical language serves both as a scalable source of supervision and as a flexible interface for interacting with complex physiological data.

A central implication of this work is that clinical reports can provide a fundamentally different form of supervision for medical AI. Conventional EEG models, including SCORE-AI^1^ and MORGOTH^2^, have achieved impressive diagnostic performance through extensive expert annotations for predefined tasks: SCORE-AI was developed on recordings annotated by 17 experts, and MORGOTH’s training sets for rhythmic and periodic patterns, interictal discharges and sleep staging required 124, 24 and 7 experts, respectively^1,2^. Although highly effective, this approach is difficult to scale across the full spectrum of clinical findings, as expanding diagnostic capabilities typically requires additional annotations and task-specific model development. Routine clinical reports offer an alternative that is both more scalable and richer in clinical information. Rather than reducing complex recordings to isolated diagnostic labels, they capture multiple findings, their spatial and temporal characteristics, relationships, uncertainty and clinical significance. Because such reports are generated as part of routine care, large collections of EEG–report pairs can be assembled without undertaking dedicated annotation campaigns for each diagnostic task. Related approaches have begun to explore report-based supervision in computational pathology and EEG–language modeling^3,4^. Our findings extend this principle by demonstrating that learning from routine clinical documentation can support competitive diagnostic performance together with broad, language-based clinical interpretation.

Importantly, this increased flexibility did not come at the expense of diagnostic performance on the evaluated shared tasks. EEG Copilot achieved virtually identical aggregate discrimination to MORGOTH across two external cohorts, despite the latter having been developed using extensive dedicated expert annotations. Comparable results against direct neurologist review of the underlying EEG recordings further corroborated this finding. This is particularly consequential because the annotation requirements of conventional approaches have been a major constraint on developing EEG models that encompass the diversity of clinical neurophysiology. Our results demonstrate that competitive diagnostic performance can be achieved using clinical reports as the primary source of task supervision, rather than relying on dedicated expert annotations for each finding. The implications extend beyond reducing annotation effort: they suggest that the information contained in routine clinical documentation can support models that learn more generalizable relationships between complex physiological signals and their clinical interpretation.

A second major advance is the breadth and flexibility of diagnostic assessment enabled by language conditioning. Whereas conventional classification-based systems are largely restricted to predefined output categories, EEG Copilot can be questioned directly about diverse EEG characteristics without requiring a separate prediction head for each finding. We demonstrated this capability across 23 clinical characteristics, substantially extending the repertoire assessed in the direct comparison with MORGOTH. Crucially, this breadth was achieved within a single model and across

English-and German-language clinical settings, with consistent aggregate performance across four hospital cohorts. This changes the interaction between clinicians and automated EEG analysis: rather than being limited to the categories anticipated during model development, clinicians can formulate diagnostic questions in the language of clinical practice. Although additional questions require individual validation, the language-based architecture offers an extensible interface through which new clinical concepts can be assessed without developing a dedicated classifier for each one. Such flexibility is particularly relevant to EEG, where clinically meaningful distinctions frequently extend beyond a limited set of diagnostic categories.

Beyond answering questions about individual abnormalities, EEG Copilot demonstrated the ability to synthesize information into integrated clinical impressions. This represents an important step toward aligning automated EEG analysis with clinical practice, where interpretation involves not only identifying patterns but also describing their morphology, distribution, relationships and significance in a coherent narrative. Across four hospital cohorts, most generated impressions were judged correct or clinically usable with minor errors by a neurologist, while substantially greater agreement with matched than shuffled reference reports indicated that the generated text incorporated recording-specific clinical information. These capabilities distinguish language-based interpretation from conventional systems that communicate their findings primarily through separate classification scores. They also create opportunities for a more interactive form of clinical decision support, in which physicians can request focused assessments, examine different aspects of a recording and use generated impressions as a starting point for clinical reporting. Further improvement in completeness and reliability will be essential, but the ability to express complex EEG information directly in medical language establishes a foundation for AI assistance that fits more naturally into clinical interpretation workflows.

The evaluation strategy is particularly important for interpreting these advances. In contrast to classification models with clearly defined reference labels, generative medical AI presents substantial challenges for objective validation. Clinical reports are imperfect references, reflecting differences in expertise, terminology, documentation practices and the findings considered relevant by individual readers. Indeed, direct expert review revealed meaningful discrepancies between original clinical reports and the underlying EEG findings, illustrating the limitations of treating routine documentation as an unquestioned ground truth. Similarly, agreement between a generated and reference impression does not necessarily establish that the model has interpreted the recording correctly, particularly because standardized clinical descriptions may receive favorable ratings even when unrelated to the underlying EEG. We therefore combined large-scale report-based evaluations with direct neurologist review of EEG recordings, comparison against an established expert-supervised model, assessment of generated impressions and shuffled-report controls. This complementary evaluation provides stronger evidence than any individual metric and highlights the importance of distinguishing linguistic agreement, diagnostic discrimination and clinically meaningful interpretation when assessing medical language models.

Several findings also identify opportunities to improve this approach. Diagnostic discrimination was generally stronger for widespread or sustained abnormalities, such as generalized slowing, than for brief or spatially restricted findings, including epileptiform discharges and seizures. One possible explanation is the challenge of preserving temporally localized information when compressing long multichannel recordings into a compact representation, compounded by the limited temporal specificity of routine clinical reports. Moreover, generating an integrated impression requires not only recognizing findings but also selecting and communicating clinically relevant information; consequently, abnormalities that can be identified through targeted questions may still be omitted from narrative reports. These observations suggest several complementary directions for further development, including explicit localization of EEG evidence, tighter alignment between findings and their temporal context, and improved report generation through structured clinical feedback. Incorporating higher-quality expert-reviewed reports and structured EEG documentation may further improve performance, particularly for uncommon or subtle abnormalities. Ultimately, combining the scalability of routine report-based learning with targeted expert supervision may offer a particularly effective strategy for further improving clinical reliability.

Several limitations should be considered. The direct EEG-based expert evaluation was restricted to 200 recordings assessed by a single neurologist, while assessments of generated impressions relied on text-based comparisons rather than independent review of the corresponding EEG signals. Large-scale diagnostic evaluations used report-derived reference labels, which are vulnerable to omissions and variation in reporting practices, and the observed similarity between models does not establish formal statistical equivalence. Although performance was demonstrated in English-and German-language settings, language and hospital site were confounded, preventing isolation of language-specific effects. Model development was restricted to adult EEG recordings with a standardized channel configuration and without additional clinical context, and the accuracy of unrestricted responses to arbitrary questions and multi-turn clinical interactions remains to be established. Most importantly, the study did not assess prospective use in clinical workflows or its effects on physician performance, diagnostic decisions or patient outcomes. Future studies should therefore investigate human–AI interaction, clinical safety and reliability, and the benefits of integrating EEG-grounded explanations and patient-specific context into physician-supervised interpretation.

Together, these findings demonstrate that the development of medical AI need not be constrained to predefined diagnostic labels and the expert annotation effort required to create them. By learning from the clinical interpretations already generated during routine care, EEG Copilot combines competitive diagnostic performance with broader, multilingual and narrative capabilities that more closely reflect how physicians interrogate and communicate complex physiological information. Beyond EEG, this approach offers a path toward medical AI systems in which clinical language provides both the foundation for scalable learning and the interface through which clinicians interact with their data.

## Methods

### Study design and datasets

We developed EEG Copilot using clinical EEG recordings and reports from Massachusetts General Hospital and Brigham and Women’s Hospital^5^ and evaluated it in external cohorts from Charité – Universitätsmedizin Berlin. The retrospective study was conducted under institutional review board approvals from Mass General Brigham (2013P001024) and the Ethics Committee of Charité – Universitätsmedizin Berlin (EA4/073/26), with informed consent waived at both institutions. Separate analyses assessed frozen signal representations, EEG–language alignment, structured question answering, and impression generation. Report-derived reference labels were distinguished from expert assessments based on direct EEG review.

Model development used EEG recordings from the two Boston sites represented in the Brain Data Science Platform dataset^5^. Report selection targeted examinations labeled “Routine” in the source metadata and patients aged older than 18 years. The signal archive contained 36,450 EDF recordings. Preprocessing excluded 59 recordings without valid windows and two recordings with insufficient canonical EEG channels, leaving 36,389 recordings from 29,958 distinct dataset patient identifiers.

Patients were randomly assigned to training, validation, and test partitions in proportions of 80%, 10%, and 10%, respectively, using a fixed seed of 42. All recordings and windows associated with a patient identifier retained the same assignment. After preprocessing, the partitions contained 29,050 training, 3,630 validation, and 3,709 test recordings.

Self-supervised encoder training used only the Boston training partition, comprising 2,283,348 retained 30-s windows. Consequently, patients assigned to the internal validation and test partitions did not contribute recordings to encoder pretraining. Subsequent EEG–language training retained the same patient assignments and applied additional requirements concerning recording duration and availability of matched text. Bridge training included 28,321 recordings. The final instruction-training mixture included 26,565 recordings, of which 26,301 supplied standalone impressions and focused question–answer examples.

External evaluation used recordings from Charité Campus Mitte. Of 2,639 available EDF recordings, 2,591 retained valid windows after preprocessing; five recordings lacked sufficient canonical EEG channels and 43 yielded no valid windows. Following duration filtering and linkage to reports and extracted labels, 2,163 recordings entered the external evaluation. Charité recordings were not used in the inspected encoder, bridge, or language-model training stages. The external archive’s storage partitions were pooled for evaluation rather than used as training and test partitions.

### Signal preparation

EDF recordings were loaded using MNE python (v1.12.1) ^11,12^. Channel names were standardized to a canonical set of 19 electrodes: Fp1, Fp2, F7, F3, Fz, F4, F8, T3, C3, Cz, C4, T4, T5, P3, Pz, P4, T6, O1, and O2. Modern temporal-channel names T7, T8, P7, and P8 were mapped to T3, T4, T5, and T6, respectively. Recordings were required to contain all 19 canonical EEG channels.

Selected EEG and ECG channels were jointly resampled to 256 Hz when their original sampling frequency differed. EEG signals were referenced to the instantaneous average across the selected EEG electrodes; ECG and other auxiliary channels were excluded from the reference. Signals were converted to microvolts and stored without per-channel amplitude normalization. When separate left and right ECG channels were available, their difference was retained; otherwise, the first identified ECG/EKG channel was used. Missing ECG was represented by zeros and an explicit availability mask.

Signals were divided into non-overlapping 30-s windows containing 7,680 samples. Incomplete terminal windows were discarded. A window was rejected if any selected EEG value was nonfinite, if any EEG channel had a standard deviation below 0.01 µV after preprocessing, or if an available ECG contained nonfinite values. Recordings without any retained windows were excluded. No additional explicit notch or band-pass filtering, independent-component artifact removal, or amplitude-threshold rejection was applied.

For recording-level EEG–language modeling, duration eligibility was calculated as the interval from the first retained window’s start to the last retained window’s end. Recordings exceeding two hours under this definition were excluded. For recording-level question answering and impression generation, the model received all retained 30-second windows from each eligible recording, without temporal cropping. Within the encoder, each channel was zero-centered within each window; EEG signals were subsequently scaled by their joint root-mean-square amplitude, while ECG was scaled separately.

### Self-supervised EEG encoder

EEG2Vec combined a temporal convolutional encoder with a six-layer transformer of hidden dimension 512 and eight attention heads. Convolutional weights were shared across EEG channels; ECG used a separate convolutional encoder. The temporal stride was 128 samples, yielding 60 tokens per channel for each 30-s window. Tokens received sinusoidal temporal embeddings and learned embeddings describing channel identity, montage view, lead type, and source and reference electrodes. A summary token and amplitude tokens were included in the transformer input.

Pretraining used paired referential and bipolar views derived from the same recording. Batches alternated between a full montage containing 19 referential and 18 longitudinal bipolar leads and a reduced montage containing six referential and four bipolar leads.

An exponential-moving-average teacher, conceptually related to data2vec^13^ received unmasked inputs, while the student received inputs with randomly masked channels and temporal token spans. The configured masking procedure hid two to four valid channels and two to three temporal spans of three tokens, aligned across channels. Temporal reconstruction loss was evaluated at span centers; masked-channel reconstruction was evaluated across the affected channel tokens.

The objective combined latent-token reconstruction, relative channel-amplitude prediction, prediction of the pooled teacher representation from the student summary token, variance and covariance regularization, and cross-view consistency:

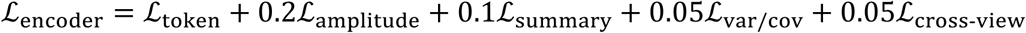

Cross-view consistency minimized the cosine distance between summary representations obtained from referential and bipolar EEG views. Teacher parameters were updated with momentum 0.999.

### Probing frozen EEG representations

We assessed information recoverable from the encoder’s 512-dimensional window summary vectors using independent classifiers for each clinical feature. The linear probe applied a single linear output layer to the mean of all retained window vectors. The recurrent probe used a bidirectional, single-layer long short-term memory network with 64 hidden units per direction, followed by concatenated mean and maximum pooling, dropout of 0.1, and a linear output layer. Recurrent state was reset across gaps exceeding 30 s.

Probes were trained with class-balanced binary cross-entropy using AdamW^14^, a learning rate of 10^−3^, weight decay of 10^−4^, and batches of 64 recordings. Training continued for at most 100 epochs, with early stopping after ten epochs without sufficient improvement in balanced validation loss. The checkpoint with the lowest balanced validation loss was restored before test evaluation. Three seeds—17, 29, and 43—were evaluated, and reported AUROCs were arithmetic means across seeds.

Unmentioned pathological findings and ECG abnormalities were treated as negative. Unmentioned sleep and normality findings, and explicitly uncertain or unassessable answers, remained unknown and were excluded for the corresponding feature. Figure 2e presents 25 EEG/ECG endpoints, excluding clinical-history questions. Confidence intervals were obtained from 1,000 patient-cluster bootstrap resamples, using common patient draws across architectures and seeds within each feature.

### Hierarchical EEG–language bridge

A local attention module compressed the frozen encoder’s patch tokens into eight learned query tokens per window^7^. These were concatenated with a projected window summary token, producing nine 768-dimensional tokens per window. Learned window-position embeddings encoded their order. A global query transformer initialized from BiomedBERT^15^ used 64 learned queries and cross-attention after every second transformer layer to aggregate the recording into 64 output tokens. Bridge pretraining combined four objectives inspired by language-image pre training^16^: bidirectional report-level contrastive alignment, statement-level contrastive ranking, joint EEG–statement matching, and EEG-conditioned text generation. The report-level objective treated texts associated with the same recording as positive matches. Statement objectives ranked report-consistent descriptions above counterfactual alternatives. EEG–text similarity was calculated using the maximum cosine similarity across query tokens. The combined loss was

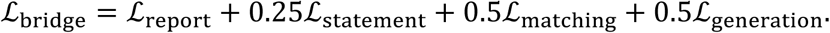

Validation monitored these objectives, statement-selection accuracy, teacher-forced next-token accuracy, and bidirectional retrieval within candidate batches. Retrieval accuracy therefore reflected the evaluated batch composition rather than unrestricted retrieval across the full dataset.

### Instruction data and language-model adaptation

Clinical reports were transformed into English-and German-language instruction examples comprising standalone impressions, focused question–answer pairs, and multi-turn dialogues. Qwen3.8-27B^17,18^ generated one impression and five focused question–answer pairs per language and report. Derived examples retained their source recording’s dataset assignment. Clinical reports supplied training targets; they were not provided to EEG Copilot during standalone question answering or impression generation.

The bridge’s 64 output tokens were projected into the 3,840-dimensional embedding space of Gemma 4 12B^19^ using layer normalization and a two-layer projection with an intermediate dimension of 2,048, SiLU activation^20^, and dropout of 0.05. The projected tokens were inserted as a continuous EEG prefix. Training minimized next-token cross-entropy over assistant responses, excluding user and system tokens from supervision.

The EEG encoder and base language-model weights remained frozen. Training progressively activated the projection, low-rank language-model adapters, query-transformer cross-attention, query-transformer backbone, and local compressor. Adapter training began at update 1,500, cross-attention at 9,000, backbone adaptation at 15,000, and local-compressor adaptation at 21,000. Low-rank adapters^6^ had rank and scaling factor 128, dropout of 0.1, and targeted the language model’s query, key, value, and output projections.

The final continuation, from update 108,000 to 118,000, sampled impressions, focused questions, and multi-turn dialogues in proportions of 50%, 30%, and 20%, respectively. Recordings were sampled uniformly within each source. Impression and focused-question losses received recording weights based on positive pathological findings: each finding’s boost was its inverse square-root training prevalence, capped at four; recordings received the maximum applicable boost or a baseline of one. Weights were normalized to a mean of one within each source. Multi-turn losses and evaluation metrics remained unweighted. An auxiliary classifier was trained on detached bridge representations and did not update the generative model.

Final model training used four H200 GPUs, bfloat16 mixed precision, and an effective batch size of 64 examples. AdamW^14^ learning rates were 2 × 10^−4^ for the projection, 2 × 10^−5^ for adapters, 10^−5^ for cross-attention, and 5 × 10^−6^ for the backbone and local compressor.

### Assessment of EEG conditioning

We compared reference-response negative log-likelihood under matched and shuffled EEG inputs while holding the text context and target response constant:

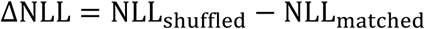

Positive values indicate that the matched recording increases the likelihood of the reference response. Because monitoring settings changed across training restarts, the concatenated trajectory was interpreted as a descriptive training diagnostic rather than a controlled comparison between training stages.

### Structured question answering

We evaluated 25 binary questions covering epileptiform activity, slowing and encephalopathy, periodic and rhythmic patterns, sleep, ECG abnormalities, and overall normality. The questions were based on the SCORE-EEG system for structured reporting of EEGs^21^ and the ACNS criteria^22^. Reference labels were extracted from clinical reports using MedGemma 27B^23^. Each question was presented independently with the EEG representation, without the report or preceding reference answers.

For the internal test analysis shown in Figure 4, the prompt specified to only answer with “Yes” or “No”, and the score compared the likelihoods of these tokens:

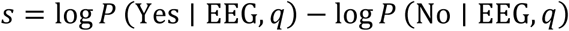

The external German analysis analogously compared “Ja” and “Nein.” Candidate scores summed conditional log probabilities over the complete answer text, excluding the turn terminator and without length normalization. This distinction matters because “Nein” comprises two tokens with the saved tokenizer.

Feature discrimination was summarized by AUROC. Category summaries were unweighted means of feature AUROCs.

### Impression generation and assessment

Impressions were generated from the EEG and a fixed request in each site’s reporting language (English for MGH and BWH, German for CBF and CCM), using greedy decoding with up to 512 new tokens and without access to the report. References were the impression sections of the original reports, used without translation or rewriting.

Qwen3.8-27B^17,18^ rated each pair, without the EEG or model identity, for correctness of the main interpretation, completeness of important reference findings and specificity of generated claims, each on a 0–2 scale using a fixed rubric. Pairs with inadequate references were excluded, leaving 2,378 MGH, 973 BWH, 3,830 CBF and 2,151 CCM pairs. As a chance baseline, the assessment was repeated after five within-site permutations of generated impressions, excluding same-recording and same-patient pairs.

A board-certified neurologist specialized in EEG and epilepsy reviewed 200 randomly sampled impressions per site against the reference text, without inspecting the EEG. Individual statements were assigned to seven clinical categories and rated fully correct (2), partly correct (1) or wrong (0); omissions and unsupported additions (fabrications) were annotated separately. Finally, each impression was rated as correct, clinically usable with minor errors or clinically unusable. Statement-level denominators excluded omissions, omission counts were not converted into rates, and 95% confidence intervals were calculated with the Wilson method.

### External expert evaluation

External expert evaluation comprised 200 randomly sampled EEG recordings acquired at Charité–Universitätsmedizin Berlin between 2023 and 2025, including examinations from general inpatient wards, intensive care units, and outpatient settings. Each recording was manually reviewed by a board-certified neurologist specialized in EEG and epilepsy. The reviewer assessed 25 SCORE-EEG-based items, recording each finding as present (“yes”), absent (“no”), or unassessable (“N/A”). These recording-based assessments served as the expert reference for evaluation of the original clinical reports.

The reviewer subsequently assessed each original report against the findings established through direct EEG review. Text segments were assigned to the corresponding SCORE-EEG-based items and classified as correct, containing a minor error, or wrong. Findings identified during EEG review but omitted from the original report were documented separately.

Each report also received an overall rating of “correct,” “clinically usable with minor errors,” or “wrong/clinically not usable.” Reports were classified as clinically unusable if their overall interpretation as normal or abnormal disagreed with the expert assessment, or if they omitted or falsely asserted clinically important findings with implications for treatment, such as epileptiform activity or burst suppression.

EEG Copilot answers and MORGOTH predictions were matched to the expert assessments by recording and corresponding clinical finding. The findings established through direct EEG review served as the reference for all comparisons; original clinical reports were evaluated as a separate comparator rather than treated as ground truth. Comparisons with Morgoth were restricted to endpoints shared by the two systems and the expert assessment. Analyses were performed separately for each finding using recordings with an assessable expert reference and an evaluable corresponding output.

For binary findings, performance was summarized by balanced accuracy, calculated as the arithmetic mean of sensitivity and specificity. Agreement with the expert reference was additionally quantified using Cohen’s κ^24^ and Gwet’s AC1^25^. Both coefficients were reported to characterize agreement under the different prevalence distributions of the evaluated findings. The numbers of reference-positive and reference-negative recordings were reported for each endpoint. Numeric estimates of posterior dominant rhythm frequency, background frequency, and posterior dominant rhythm amplitude were assessed separately using range overlap.

Summary agreement coefficients were calculated as unweighted means across eligible findings. To limit the contribution of sparsely represented endpoints, these summaries included only findings with at least ten reference-positive and ten reference-negative recordings. The comparison across all shared findings and the comparison restricted to endpoints available from MORGOTH used their respective eligible endpoint sets. Uncertainty in the mean agreement coefficients was summarized using 95% bootstrap confidence intervals. Comparisons between systems were descriptive; differences in point estimates were not interpreted as evidence of statistically significant differences.

## Data availability

The EEG recordings and associated clinical reports from Massachusetts General Hospital (MGH) and Brigham and Women’s Hospital (BWH) are available through the Harvard Electroencephalography Database^5^, hosted by the Brain Data Science Platform (BDSP; https://bdsp.io/content/harvard-eeg-db/4.1/). Access is subject to credential verification and a data use agreement. The clinical EEG recordings and associated reports from Charité – Universitätsmedizin Berlin are not publicly available owing to patient privacy, data protection regulations and institutional restrictions on sharing patient-level clinical data.

## Code availability

Selected code developed for this study, including EEG preprocessing and evaluation utilities, will be made publicly available upon publication through the Computational Neurology GitLab repository (https://gitlab.com/computational-neurologie). Publicly accessible third-party resources used in this study include MNE-Python^11,12^, BiomedBERT^15^, Gemma 4^19^, Qwen3.8-27B^17,18^, MedGemma 27B^23^ and MORGOTH^2^, which are available through their respective official repositories and subject to applicable licensing conditions.

## Supplementary Material

### Results Charité Campus Mitte (CCM)

**Table 1.** Detailed comparison metrics for findings supported by both MORGOTH and the EEG Copilot on the CCM cohort.

| **Metric** | **Morgoth** | **EEG Copilot** |
|---|---|---|
| **AUROC** | 0.796<br>(0.777–0.813) | 0.802<br>(0.782–0.822) |
| **AP** | 0.352<br>(0.330–0.381) | 0.353<br>(0.332–0.386) |
| **Acc.** | 0.832<br>(0.827–0.838) | 0.810<br>(0.803–0.818) |
| **BA** | 0.709<br>(0.688–0.729) | 0.705<br>(0.687–0.724) |
| **Sens.** | 0.529<br>(0.487–0.568) | 0.605<br>(0.569–0.642) |
| **Spec.** | 0.890<br>(0.884–0.895) | 0.805<br>(0.796–0.813) |
| **PPV** | 0.396<br>(0.374–0.419) | 0.341<br>(0.296–0.390) |
| **F1** | 0.357<br>(0.331–0.382) | 0.367<br>(0.337–0.395) |
| **Gwet’s AC1** | 0.730<br>(0.720–0.741) | 0.698<br>(0.683–0.712) |
| **Cohen’s κ** | 0.282<br>(0.257–0.307) | 0.282<br>(0.251–0.311) |

### Results Charité Benjamin Franklin (CBF)

**Table 2.** Detailed comparison metrics for findings supported by both MORGOTH and the EEG Copilot on the CBF cohort.

| **Metric** | **Morgoth** | **EEG Copilot** |
|---|---|---|
| **AUROC** | 0.811<br>(0.802–0.821) | 0.813<br>(0.801–0.824) |
| **AP** | 0.395<br>(0.381–0.412) | 0.404<br>(0.385–0.430) |
| **Acc.** | 0.822<br>(0.818–0.827) | 0.804<br>(0.800–0.809) |
| **BA** | 0.744<br>(0.734–0.754) | 0.679<br>(0.671–0.688) |
| **Sens.** | 0.624<br>(0.604–0.643) | 0.537<br>(0.520–0.555) |
| **Spec.** | 0.864<br>(0.860–0.869) | 0.821<br>(0.816–0.827) |
| **PPV** | 0.367<br>(0.354–0.381) | 0.384<br>(0.354–0.415) |
| **F1** | 0.384<br>(0.369–0.398) | 0.351<br>(0.333–0.368) |
| **Gwet’s AC1** | 0.729<br>(0.722–0.736) | 0.684<br>(0.676–0.692) |
| **Cohen’s κ** | 0.297<br>(0.282–0.312) | 0.257<br>(0.239–0.274) |

### Expert reference cohort

**Table 3.** Detailed comparison metrics for findings supported by both MORGOTH and the EEG Copilot on the expert reference cohort.

| **Metric** | **Morgoth** | **EEG Copilot** |
|---|---|---|
| **AUROC** | 0.835<br>(0.803–0.865) | 0.861<br>(0.825–0.893) |
| **AP** | 0.531<br>(0.473–0.629) | 0.501<br>(0.414–0.610) |
| **Acc.** | 0.824<br>(0.808–0.839) | 0.813<br>(0.792–0.833) |
| **BA** | 0.746<br>(0.714–0.776) | 0.744<br>(0.684–0.793) |
| **Sens.** | 0.588<br>(0.524–0.647) | 0.691<br>(0.572–0.786) |
| **Spec.** | 0.904<br>(0.891–0.916) | 0.797<br>(0.771–0.823) |
| **PPV** | 0.537<br>(0.434–0.630) | 0.417<br>(0.248–0.497) |
| **F1** | 0.411<br>(0.303–0.486) | 0.450<br>(0.314–0.516) |
| **Gwet’s AC1** | 0.538<br>(0.490–0.582) | 0.614<br>(0.564–0.664) |
| **Cohen’s κ** | 0.244<br>(0.185–0.295) | 0.383<br>(0.319–0.443) |

**Table 4.** Detailed metrics for the ǪA analysis on the MGH cohort.

| Finding | Ns (n+) | Ng (n+) | AUROC | AP | Acc. | BA | Sens. | Spec. | PPV | F1 |
|---|---|---|---|---|---|---|---|---|---|---|
| Normal EEG | 2557 (1021) | 2556 (1021) | 0.857 | 0.736 | 0.773 | 0.786 | 0.853 | 0.720 | 0.669 | 0.750 |
| Abnormal EEG | 2557 (1548) | 2073 (1136) | 0.852 | 0.914 | 0.756 | 0.775 | 0.577 | 0.974 | 0.965 | 0.722 |
| Drowsiness | 2267 (1880) | 2249 (1869) | 0.788 | 0.949 | 0.712 | 0.710 | 0.713 | 0.708 | 0.923 | 0.804 |
| Sleep | 2438 (937) | 1200 (652) | 0.885 | 0.851 | 0.838 | 0.837 | 0.854 | 0.819 | 0.849 | 0.852 |
| K-complexes | 801 (680) | 801 (680) | 0.937 | 0.986 | 0.888 | 0.866 | 0.897 | 0.835 | 0.968 | 0.931 |
| Sleep spindles | 1464 (823) | 1461 (822) | 0.897 | 0.914 | 0.828 | 0.825 | 0.849 | 0.801 | 0.846 | 0.848 |
| Vertex waves | 866 (771) | 866 (771) | 0.891 | 0.985 | 0.844 | 0.792 | 0.859 | 0.726 | 0.962 | 0.907 |
| Focal slowing | 2557 (896) | 2557 (896) | 0.700 | 0.588 | 0.709 | 0.633 | 0.377 | 0.889 | 0.646 | 0.476 |
| Generalized slowing | 2557 (1017) | 2557 (1017) | 0.926 | 0.904 | 0.844 | 0.853 | 0.898 | 0.809 | 0.756 | 0.821 |
| Burst suppression | 2557 (38) | 2557 (38) | 0.899 | 0.171 | 0.985 | 0.500 | 0.000 | 1.000 | 0.000 | 0.000 |
| GRDA | 2557 (193) | 2557 (193) | 0.810 | 0.279 | 0.925 | 0.500 | 0.000 | 1.000 | n.a. | 0.000 |
| LRDA | 2557 (130) | 2557 (130) | 0.710 | 0.132 | 0.948 | 0.518 | 0.038 | 0.997 | 0.417 | 0.070 |
| Pathological rhythmic activity | 2557 (1343) | 2557 (1343) | 0.781 | 0.792 | 0.616 | 0.632 | 0.321 | 0.942 | 0.860 | 0.467 |
| BiPD | 2557 (12) | 2557 (12) | 0.857 | 0.019 | 0.603 | 0.801 | 1.000 | 0.601 | 0.012 | 0.023 |
| BIRDs | 2557 (76) | 2557 (76) | 0.698 | 0.101 | 0.741 | 0.624 | 0.500 | 0.749 | 0.057 | 0.103 |
| GPD | 2557 (132) | 2557 (132) | 0.831 | 0.312 | 0.860 | 0.754 | 0.636 | 0.872 | 0.213 | 0.319 |
| LPD | 2557 (95) | 2557 (95) | 0.675 | 0.072 | 0.949 | 0.528 | 0.074 | 0.983 | 0.143 | 0.097 |
| Periodic discharges | 2557 (274) | 2557 (274) | 0.844 | 0.441 | 0.876 | 0.694 | 0.464 | 0.925 | 0.426 | 0.444 |
| SIPD | 2557 (18) | 2557 (18) | 0.857 | 0.033 | 0.628 | 0.813 | 1.000 | 0.625 | 0.019 | 0.036 |
| Triphasic waves | 2557 (114) | 2557 (114) | 0.897 | 0.258 | 0.435 | 0.700 | 0.991 | 0.409 | 0.073 | 0.135 |
| Seizure | 2557 (46) | 2551 (45) | 0.679 | 0.066 | 0.982 | 0.500 | 0.000 | 1.000 | 0.000 | 0.000 |
| Status epilepticus | 2557 (30) | 2557 (30) | 0.897 | 0.174 | 0.922 | 0.779 | 0.633 | 0.925 | 0.091 | 0.160 |
| Interictal discharges | 2557 (557) | 2557 (557) | 0.745 | 0.476 | 0.795 | 0.630 | 0.338 | 0.922 | 0.547 | 0.417 |

**Table 5.** Detailed metrics for the ǪA analysis on the BWH cohort.

| Finding | Ns (n+) | Ng (n+) | AUROC | AP | Acc. | BA | Sens. | Spec. | PPV | F1 |
|---|---|---|---|---|---|---|---|---|---|---|
| Normal EEG | 1054 (274) | 1054 (274) | 0.833 | 0.584 | 0.694 | 0.760 | 0.898 | 0.622 | 0.455 | 0.604 |
| Abnormal EEG | 1054 (768) | 1011 (730) | 0.838 | 0.936 | 0.727 | 0.768 | 0.675 | 0.861 | 0.927 | 0.781 |
| Drowsiness | 1022 (953) | 1020 (952) | 0.831 | 0.985 | 0.794 | 0.699 | 0.809 | 0.588 | 0.965 | 0.880 |
| Sleep | 1040 (435) | 638 (319) | 0.830 | 0.817 | 0.806 | 0.806 | 0.768 | 0.843 | 0.831 | 0.798 |
| K-complexes | 222 (211) | 222 (211) | 0.841 | 0.987 | 0.883 | 0.766 | 0.896 | 0.636 | 0.979 | 0.936 |
| Sleep spindles | 809 (396) | 809 (396) | 0.849 | 0.852 | 0.771 | 0.771 | 0.775 | 0.768 | 0.762 | 0.768 |
| Vertex waves | 404 (386) | 404 (386) | 0.822 | 0.991 | 0.814 | 0.691 | 0.826 | 0.556 | 0.976 | 0.895 |
| Focal slowing | 1054 (563) | 1054 (563) | 0.689 | 0.711 | 0.638 | 0.642 | 0.581 | 0.703 | 0.691 | 0.631 |
| Generalized slowing | 1054 (472) | 1054 (472) | 0.898 | 0.888 | 0.821 | 0.821 | 0.828 | 0.814 | 0.784 | 0.805 |
| Burst suppression | 1054 (16) | 1054 (16) | 0.926 | 0.173 | 0.984 | 0.500 | 0.000 | 0.999 | 0.000 | 0.000 |
| GRDA | 1054 (103) | 1054 (103) | 0.802 | 0.312 | 0.903 | 0.505 | 0.010 | 1.000 | 1.000 | 0.019 |
| LRDA | 1054 (59) | 1054 (59) | 0.677 | 0.140 | 0.900 | 0.581 | 0.220 | 0.941 | 0.181 | 0.198 |
| Pathological rhythmic activity | 1054 (562) | 1054 (562) | 0.739 | 0.777 | 0.655 | 0.666 | 0.500 | 0.831 | 0.772 | 0.607 |
| BiPD | 1054 (6) | 1054 (6) | 0.868 | 0.076 | 0.446 | 0.721 | 1.000 | 0.443 | 0.010 | 0.020 |
| BIRDs | 1054 (16) | 1054 (16) | 0.717 | 0.041 | 0.796 | 0.650 | 0.500 | 0.801 | 0.037 | 0.069 |
| GPD | 1054 (70) | 1054 (70) | 0.910 | 0.539 | 0.851 | 0.827 | 0.800 | 0.855 | 0.281 | 0.416 |
| LPD | 1054 (56) | 1054 (56) | 0.666 | 0.106 | 0.875 | 0.588 | 0.268 | 0.909 | 0.142 | 0.185 |
| Periodic discharges | 1054 (143) | 1054 (143) | 0.856 | 0.580 | 0.875 | 0.739 | 0.552 | 0.925 | 0.537 | 0.545 |
| SIPD | 1054 (5) | 1054 (5) | 0.812 | 0.044 | 0.480 | 0.639 | 0.800 | 0.479 | 0.007 | 0.014 |
| Triphasic waves | 1054 (89) | 1054 (89) | 0.923 | 0.536 | 0.380 | 0.661 | 1.000 | 0.322 | 0.120 | 0.214 |
| Seizure | 1054 (12) | 1054 (12) | 0.647 | 0.034 | 0.983 | 0.497 | 0.000 | 0.994 | 0.000 | 0.000 |
| Status epilepticus | 1054 (10) | 1054 (10) | 0.769 | 0.040 | 0.921 | 0.663 | 0.400 | 0.926 | 0.049 | 0.088 |
| Interictal discharges | 1054 (225) | 1054 (225) | 0.676 | 0.352 | 0.725 | 0.600 | 0.382 | 0.818 | 0.363 | 0.372 |

**Table 6.** Detailed metrics for the ǪA analysis on the CBF cohort.

| Finding | Ns (n+) | Ng (n+) | AUROC | AP | Acc. | BA | Sens. | Spec. | PPV | F1 |
|---|---|---|---|---|---|---|---|---|---|---|
| Normal EEG | 3915 (1207) | 2196 (412) | 0.872 | 0.671 | 0.875 | 0.874 | 0.874 | 0.875 | 0.617 | 0.724 |
| Abnormal EEG | 3915 (2987) | 3726 (2890) | 0.868 | 0.955 | 0.806 | 0.811 | 0.802 | 0.821 | 0.939 | 0.865 |
| Drowsiness | 3030 (1525) | 3028 (1525) | 0.610 | 0.632 | 0.570 | 0.569 | 0.729 | 0.408 | 0.555 | 0.631 |
| Sleep | 2953 (140) | 2699 (137) | 0.856 | 0.379 | 0.840 | 0.771 | 0.693 | 0.848 | 0.196 | 0.306 |
| K-complexes | 3820 (96) | 3820 (96) | 0.784 | 0.205 | 0.658 | 0.693 | 0.729 | 0.656 | 0.052 | 0.097 |
| Sleep spindles | 579 (79) | 577 (79) | 0.940 | 0.735 | 0.901 | 0.842 | 0.759 | 0.924 | 0.612 | 0.678 |
| Vertex waves | 3855 (71) | 3855 (71) | 0.891 | 0.190 | 0.608 | 0.766 | 0.930 | 0.602 | 0.042 | 0.080 |
| Focal slowing | 3914 (2274) | 3914 (2274) | 0.704 | 0.766 | 0.607 | 0.631 | 0.485 | 0.777 | 0.751 | 0.589 |
| Generalized slowing | 3915 (2052) | 3915 (2052) | 0.925 | 0.929 | 0.822 | 0.817 | 0.934 | 0.699 | 0.774 | 0.846 |
| Burst suppression | 3915 (127) | 3915 (127) | 0.837 | 0.204 | 0.966 | 0.534 | 0.071 | 0.996 | 0.391 | 0.120 |
| GRDA | 3915 (220) | 3915 (220) | 0.800 | 0.178 | 0.601 | 0.731 | 0.877 | 0.585 | 0.112 | 0.198 |
| LRDA | 3915 (331) | 3915 (331) | 0.713 | 0.210 | 0.829 | 0.615 | 0.356 | 0.873 | 0.206 | 0.261 |
| Pathological rhythmic activity | 3915 (2482) | 3869 (2441) | 0.747 | 0.820 | 0.636 | 0.671 | 0.537 | 0.805 | 0.825 | 0.651 |
| BiPD | 3915 (9) | 3912 (9) | 0.887 | 0.025 | 0.410 | 0.704 | 1.000 | 0.408 | 0.004 | 0.008 |
| BIRDs | 3915 (147) | 3915 (147) | 0.838 | 0.175 | 0.611 | 0.755 | 0.912 | 0.599 | 0.082 | 0.150 |
| GPD | 3915 (109) | 3915 (109) | 0.851 | 0.236 | 0.509 | 0.729 | 0.963 | 0.496 | 0.052 | 0.098 |
| LPD | 3915 (44) | 3105 (37) | 0.931 | 0.286 | 0.983 | 0.537 | 0.081 | 0.993 | 0.130 | 0.100 |
| Periodic discharges | 3915 (278) | 3850 (261) | 0.872 | 0.442 | 0.926 | 0.703 | 0.444 | 0.962 | 0.457 | 0.450 |
| SIPD | 3915 (0) | 3915 (0) | n.a. | n.a. | 0.374 | n.a. | n.a. | 0.374 | 0.000 | 0.000 |
| Triphasic waves | 3915 (42) | 3915 (42) | 0.870 | 0.075 | 0.404 | 0.699 | 1.000 | 0.397 | 0.018 | 0.035 |
| Seizure | 3915 (131) | 3301 (96) | 0.686 | 0.252 | 0.973 | 0.572 | 0.146 | 0.998 | 0.667 | 0.239 |
| Status epilepticus | 3915 (156) | 3915 (156) | 0.893 | 0.296 | 0.890 | 0.764 | 0.628 | 0.901 | 0.208 | 0.313 |
| Interictal discharges | 3914 (491) | 3246 (358) | 0.762 | 0.330 | 0.842 | 0.661 | 0.430 | 0.893 | 0.332 | 0.375 |

**Table 7.** Detailed metrics for the ǪA analysis on the CCM cohort.

| **Finding** | **Ns (n+)** | **Ng (n+)** | **AUROC** | **AP** | **Acc.** | **BA** | **Sens.** | **Spec.** | **PPV** | **F1** |
|---|---|---|---|---|---|---|---|---|---|---|
| Normal EEG | 2163 (1057) | 1229 (513) | 0.883 | 0.835 | 0.895 | 0.896 | 0.899 | 0.892 | 0.857 | 0.877 |
| Abnormal EEG | 2163 (1125) | 1955 (1086) | 0.892 | 0.912 | 0.759 | 0.738 | 0.921 | 0.556 | 0.722 | 0.809 |
| Drowsiness | 846 (499) | 846 (499) | 0.518 | 0.634 | 0.537 | 0.490 | 0.747 | 0.233 | 0.584 | 0.656 |
| Sleep | 523 (76) | 502 (74) | 0.782 | 0.487 | 0.799 | 0.714 | 0.595 | 0.834 | 0.383 | 0.466 |
| K-complexes | 1792 (36) | 1792 (36) | 0.831 | 0.221 | 0.666 | 0.748 | 0.833 | 0.663 | 0.048 | 0.091 |
| Sleep spindles | 87 (22) | 87 (22) | 0.928 | 0.804 | 0.874 | 0.855 | 0.818 | 0.892 | 0.720 | 0.766 |
| Vertex waves | 2124 (47) | 2119 (47) | 0.840 | 0.228 | 0.570 | 0.718 | 0.872 | 0.563 | 0.043 | 0.082 |
| Focal slowing | 2163 (503) | 2163 (503) | 0.734 | 0.490 | 0.632 | 0.665 | 0.728 | 0.603 | 0.357 | 0.479 |
| Generalized slowing | 2163 (863) | 2163 (863) | 0.919 | 0.896 | 0.752 | 0.783 | 0.934 | 0.632 | 0.627 | 0.750 |
| Burst suppression | 2163 (37) | 2163 (37) | 0.867 | 0.266 | 0.982 | 0.540 | 0.081 | 0.998 | 0.429 | 0.136 |
| GRDA | 2163 (75) | 2163 (75) | 0.853 | 0.132 | 0.638 | 0.787 | 0.947 | 0.627 | 0.084 | 0.154 |
| LRDA | 2163 (75) | 2163 (75) | 0.795 | 0.131 | 0.831 | 0.726 | 0.613 | 0.839 | 0.120 | 0.201 |
| Pathological rhythmic activity | 2163 (788) | 2121 (766) | 0.826 | 0.741 | 0.762 | 0.747 | 0.693 | 0.801 | 0.663 | 0.678 |
| BiPD | 2163 (4) | 2162 (4) | 0.965 | 0.361 | 0.317 | 0.658 | 1.000 | 0.316 | 0.003 | 0.005 |
| BIRDs | 2163 (33) | 2163 (33) | 0.763 | 0.095 | 0.564 | 0.719 | 0.879 | 0.560 | 0.030 | 0.058 |
| GPD | 2163 (54) | 2163 (54) | 0.677 | 0.046 | 0.589 | 0.708 | 0.833 | 0.583 | 0.049 | 0.092 |
| LPD | 2163 (20) | 1800 (19) | 0.844 | 0.060 | 0.972 | 0.595 | 0.211 | 0.980 | 0.103 | 0.138 |
| Periodic discharges | 2163 (123) | 2150 (121) | 0.837 | 0.374 | 0.926 | 0.677 | 0.397 | 0.958 | 0.358 | 0.376 |
| SIPD | 2163 (2) | 2163 (2) | 0.960 | 0.017 | 0.270 | 0.635 | 1.000 | 0.269 | 0.001 | 0.003 |
| Triphasic waves | 2163 (51) | 2163 (51) | 0.910 | 0.195 | 0.396 | 0.691 | 1.000 | 0.381 | 0.038 | 0.072 |
| Seizure | 2163 (24) | 1765 (21) | 0.732 | 0.254 | 0.988 | 0.664 | 0.333 | 0.995 | 0.467 | 0.389 |
| Status epilepticus | 2163 (16) | 2163 (16) | 0.930 | 0.182 | 0.903 | 0.827 | 0.750 | 0.905 | 0.055 | 0.103 |
| Interictal discharges | 2163 (227) | 1665 (154) | 0.740 | 0.290 | 0.835 | 0.679 | 0.487 | 0.871 | 0.278 | 0.354 |

**Table 8.** Examples of original and EEG Copilot expressions chosen from correct, partially correct, and wrong subgroups.

| Original impression | Synthetic impression |
|---|---|
| This is an abnormal EEG characterized by near-continuous generalized periodic discharges (GPDs) at 1-2 Hz with a triphasic morphology and 200 uV amplitude. The background shows continuous diffuse irregular delta slowing at 0.5-2 Hz with 30-80 uV amplitude. No electrographic seizures were detected. | This EEG is abnormal due to generalized delta slowing of the background and stimulus-induced generalized periodic discharges (GPDs). The GPDs have triphasic morphology with an anterior-posterior gradient, while the accompanying rhythmic activity shows a lateralized distribution. No definite electrographic seizures were observed, though the pattern falls along the ictal-interictal continuum. |
| This is an abnormal adult EEG indicative of an encephalopathic state. It features abundant generalized periodic discharges with triphasic morphology at 0.5-2 Hz and diffuse irregular delta slowing appearing as 1-2 Hz GRDA+S. No seizures were detected. | This is an abnormal EEG characterized by a poorly organized background with diffuse delta-theta slowing and no discernible posterior dominant rhythm. Frequent generalized discharges with triphasic morphology are observed, occurring in isolation or brief runs, along with occasional 1-2 Hz GRDA. No seizures were detected. Interpretation is limited by excessive muscle artifact in the bilateral frontotemporal leads. |
| This EEG is abnormal due to abundant, fluctuating, generalized periodic discharges with triphasic morphology and generalized, polymorphic delta and theta background slowing. No seizures or interictal epileptiform discharges were detected. The ECG shows normal sinus rhythm. | This EEG is abnormal due to near-continuous 1 Hz generalized periodic discharges that are occasionally rhythmic, along with severe diffuse background slowing. The background shows irregular delta-theta activity that worsens in the second half of the recording. No electrographic seizures are observed. |
| This is an abnormal adult EEG showing occasional focal left temporal delta slowing (2-3 Hz) maximal at T5. The background is symmetric with a 9 Hz posterior dominant rhythm. No epileptiform abnormalities are present. | This is an abnormal adult EEG in the awake and drowsy states. The primary abnormality is occasional focal left temporal delta slowing (2-4 Hz). No epileptiform abnormalities or seizures were present. The background is otherwise symmetric with a 10-11 Hz posterior dominant rhythm. |
| This EEG is abnormal due to occasional focal left temporal theta and delta slowing, which is more prominent during drowsiness. These findings are consistent with moderate left temporal cerebral dysfunction. No definite epileptiform abnormalities or seizures were observed. The background consists of well-organized 9-10 Hz posterior alpha activity with good reactivity. | This routine EEG is abnormal due to intermittent left temporal theta and delta slowing, consistent with mild left temporal cerebral dysfunction. The waking background is well-organized with 9-10 Hz posterior alpha activity and good reactivity. No definite epileptiform abnormalities or seizures were identified. |
| This EEG is abnormal due to continuous diffuse irregular mixed theta and delta slowing of the background at 1-5 Hz with 30-50 uV amplitude. There were no seizures or epileptiform discharges in this recording. | This is an abnormal EEG characterized by a poorly organized background with diffuse theta and delta slowing. No seizures or epileptiform discharges were observed. The findings are consistent with moderate diffuse cerebral dysfunction or encephalopathy. |
| This is an abnormal EEG characterized by diffuse mixed theta and delta slowing of the background (2-6 Hz, 20-40 uV). A posterior dominant rhythm of 8-9 Hz (50-70 uV) is preserved with a fair anterior-posterior gradient. No epileptiform discharges or electrographic seizures were detected. The ECG shows no dysrhythmia. | This is an abnormal EEG characterized by diffuse irregular theta and delta slowing of the background. No epileptiform abnormalities or seizure activity were detected. The ECG shows no dysrhythmia. |
| This is a mildly abnormal EEG due to frequent brief runs of diffuse irregular delta-theta slowing (3-6 Hz) lasting up to 5-6 seconds. These bursts are not clearly associated with behavioral state changes. No epileptiform discharges or seizures were detected. | This portable EEG is abnormal due to frequent bilateral theta and delta slowing with shifting predominance, consistent with mild bilateral cerebral dysfunction. The waking background is well-organized with a reactive 9-10 Hz posterior dominant rhythm. No definite epileptiform abnormalities or seizures were detected. |
| The portable EEG is abnormal due to nearly-continuous, poorly-organized delta and theta slowing bilaterally with no discernible posterior dominant rhythm. These findings are consistent with moderate-to-marked bilateral cerebral dysfunction. No definite focal or epileptiform abnormalities are present. | This portable EEG performed in briefly waking and predominantly drowsy/sleeping states is abnormal due to moderate background disorganization and bilateral theta-delta slowing. These findings are consistent with moderate bilateral cerebral dysfunction and encephalopathy. There are no definite focal or epileptiform abnormalities. |
| This is an abnormal EEG characterized by continuous disorganized diffuse delta-theta slowing in the awake and drowsy states. No epileptiform abnormalities or seizure activity were detected. A significant sway artifact was present during the recording. | This portable EEG is abnormal due to a poorly organized background with bilateral theta and delta slowing, consistent with moderate bilateral cerebral dysfunction or encephalopathy. No definite focal or epileptiform abnormalities are seen. Interpretation is limited by excessive movement and myogenic artifact. |
| This is an abnormal EEG in the awake state characterized by frequent intermittent focal polymorphic delta slowing over the left hemisphere, maximal temporally. The background is symmetric with a 9-10 Hz posterior dominant rhythm and good anterior-posterior organization. No epileptiform abnormalities or electrographic seizures were detected. | This is an abnormal EEG characterized by intermittent focal theta slowing in the left temporal region. The background is otherwise symmetric with a well-organized 8-9 Hz posterior dominant rhythm. No epileptiform abnormalities or seizure activity were detected. |
| This is an abnormal EEG characterized by frequent, intermittent bitemporal polymorphic sharply contoured theta slowing (5-7 Hz, 60-80 uV) with shifting hemispheric predominance. This activity is present during both awake and drowsy states. No definite epileptiform abnormalities or electrographic seizures were detected. The background shows a symmetrical 8-9 Hz posterior dominant rhythm. | This is an abnormal EEG characterized by intermittent focal theta slowing in the left temporal region. The background is otherwise normal with a reactive 8-9 Hz posterior dominant rhythm. No epileptiform discharges or seizure activity were detected. |
| Abnormal EEG recorded in awake and drowsy states due to frequent diffuse theta and intermittent diffuse delta slowing of the background. No organized electrographic seizures were detected. Stage II sleep did not occur. | This is an abnormal EEG characterized by diffuse irregular theta and delta slowing of the background. No epileptiform abnormalities or organized electrographic seizures were detected. The recording includes periods of drowsiness and sleep with generalized slowing. |
| This is a normal EEG performed in awake and drowsy states. The background is symmetric with a reactive 8-9 Hz posterior dominant rhythm and excess beta activity. Drowsiness shows generalized slowing, but no deeper sleep was reached. No epileptiform abnormalities or seizures were detected. | This is a normal EEG in the awake and drowsy states. No epileptiform abnormalities or seizure activity were detected. The background is symmetric with a reactive 10 Hz posterior dominant rhythm, and generalized slowing was noted during drowsiness. |
| This is an abnormal EEG showing rare bifrontal or left frontal sharp waves and occasional frontal rhythmic delta slowing, most prominent on the left. The background exhibits generalized polymorphic theta slowing. No electrographic seizures were detected. | This is an abnormal EEG characterized by diffuse theta slowing and frequent generalized delta slowing. Frequent 1-2 Hz generalized rhythmic delta activity (GRDA) is present, with left-sided predominance. Occasional left frontotemporal epileptiform discharges are noted. No electrographic seizures were detected. |
| This is an abnormal EEG due to frequent left frontotemporal epileptiform discharges. Sharp waves (40-60 µV, 100 ms) are maximal at F7 with a field to T3 and T5. The background is otherwise normal with a 10 Hz posterior dominant rhythm, and typical drowsiness and sleep features are present. | This is a normal EEG in the awake, drowsy, and asleep states. No epileptiform abnormalities or seizure activity were detected. The background is symmetric with a well-organized 10 Hz posterior dominant rhythm. |
| This is a normal adult EEG in the awake state. No epileptiform abnormalities or focal/generalized slowing are present. A modest photic driving response was noted during stimulation. The accompanying ECG demonstrates bradycardia. | This is an abnormal EEG characterized by diffuse theta slowing of the background and intermittent right frontal-temporal delta slowing. Occasional right frontal-temporal epileptiform discharges are present, maximal at F4/T4. No electrographic seizures were observed. |
| Abnormal EEG due to irregular left-sided delta slowing and a left-sided rhythm. Sleep features include poorly formed vertex waves and sleep spindles. No electrographic seizures were captured. | This is an abnormal EEG characterized by diffuse irregular delta-theta slowing and intermittent focal right frontotemporal delta slowing. There is a loss of faster frequencies over the right hemisphere. No epileptiform abnormalities or seizure activity were detected. |
| This EEG is abnormal due to continuous diffuse irregular delta and theta slowing of the background. There were no seizures or epileptiform discharges in this recording. The clinical significance of the slowing is unclear in the setting of sedation. | This is an abnormal EEG characterized by a burst suppression pattern with a suppression ratio of approximately 75%. The background activity consists of bursts of 3-5 Hz theta waves (10-20 uV) alternating with periods of voltage attenuation. No epileptiform discharges or electrographic seizures were identified. |
| This is an abnormal EEG characterized by severe background suppression with symmetric low-amplitude delta activity. Continuous polyspike-and-wave discharges evolve into generalized periodic discharges time-locked with myoclonus, consistent with myoclonic status epilepticus. No drowsiness or sleep features are present. | This is an abnormal EEG characterized by diffuse irregular delta slowing of the background with absent anterior-posterior gradient. There is intermittent focal delta slowing in the right frontal region and occasional to frequent right frontal-temporal epileptiform discharges. No electrographic seizures were observed. The recording contains significant muscle and motion artifact, and the ECG channel was not recording. |

## References

1. Tveit, J. et al. Automated Interpretation of Clinical Electroencephalograms Using Artificial Intelligence. JAMA Neurol. 80, 805–812 (2023).

2. Sun, C. et al. Toward unified and comprehensive automated electroencephalogram interpretation: a multicentre development and validation of an electroencephalogram foundation model. *Lancet Digit*. Health 8, (2026).

3. Vorontsov, E. et al. End-to-end multimodal pathology foundation model with clinical dialogue. Nat. Med. 32, 3203–3213 (2026).

4. Pradeepkumar, J., Chen, Z. & Sun, J. Neural Signals Generate Clinical Notes in the Wild. Preprint at 10.48550/arXiv.2601.22197 (2026).

5. Sun, C. et al. Harvard Electroencephalography Database: A comprehensive clinical electroencephalographic resource from four Boston hospitals. Epilepsia 66, 3411–3425 (2025).

6. Hu, E. J., et al. LoRA: Low-Rank Adaptation of Large Language Models. Preprint at 10.48550/arXiv.2106.09685 (2021).

7. Vaswani, A., et al. Attention Is All You Need. Preprint at 10.48550/arXiv.1706.03762 (2023).

8. Bardes, A., Ponce, J. & LeCun, Y. VICReg: Variance-Invariance-Covariance Regularization for Self-Supervised Learning. Preprint at 10.48550/arXiv.2105.04906 (2022).

9. Huang, Z., Xu, W. & Yu, K. Bidirectional LSTM-CRF Models for Sequence Tagging. Preprint at 10.48550/arXiv.1508.01991 (2015).

10. Selivanov, A., et al. ECG-LLM: Foundation Model for ECG-Based Cardiac Reasoning. Preprint at 10.48550/arXiv.2607.16323 (2026).

11. Larson, E. et al. MNE-Python. 10.5281/zenodo.19666955(2026) doi:10.5281/zenodo.19666955.

12. Gramfort, A. et al. MEG and EEG data analysis with MNE-Python. Front. Neurosci. 7, (2013).

13. Baevski, A., et al. data2vec: A General Framework for Self-supervised Learning in Speech, Vision and Language. Preprint at 10.48550/arXiv.2202.03555 (2022).

14. Loshchilov, I. & Hutter, F. Decoupled Weight Decay Regularization. Preprint at 10.48550/arXiv.1711.05101 (2019).

15. Gu, Y. et al. Domain-Specific Language Model Pretraining for Biomedical Natural Language Processing. ACM Trans. Comput. Healthc. Health 3, 2:1–2:23 (2021).

16. Li, J., Li, D., Savarese, S. & Hoi, S. BLIP-2: Bootstrapping Language-Image Pre-training with Frozen Image Encoders and Large Language Models. Preprint at 10.48550/arXiv.2301.12597 (2023).

17. Yang, A., et al. Qwen3 Technical Report. Preprint at 10.48550/arXiv.2505.09388 (2025).

18. Team, Q. Qwen. *Qwen* https://qwenlm.github.io/blog/qwen3.8/ (2026).

19. Team, G., et al. Gemma 4 Technical Report. Preprint at 10.48550/arXiv.2607.02770 (2026).

20. Elfwing, S., Uchibe, E. & Doya, K. Sigmoid-Weighted Linear Units for Neural Network Function Approximation in Reinforcement Learning. Preprint at 10.48550/arXiv.1702.03118 (2017).

21. Beniczky, S. et al. Standardized computer-based organized reporting of EEG: SCORE – Second version. Clin. Neurophysiol. 128, 2334–2346 (2017).

22. Hirsch, L. J. et al. American Clinical Neurophysiology Society’s Standardized Critical Care EEG Terminology: 2021 Version. J. Clin. Neurophysiol. 38, 1–29 (2021).

23. Sellergren, A. et al. MedGemma Technical Report. Nat. Med. 32, 3695–3704 (2026).

24. Cohen, J. A Coefficient of Agreement for Nominal Scales. Educ. Psychol. Meas. 20, 37– 46 (1960).

25. Gwet, K. L. Computing inter-rater reliability and its variance in the presence of high agreement. Br. J. Math. Stat. Psychol. 61, 29–48 (2008).

